# The effects of time-restricted eating vs. standard dietary advice on weight, metabolic health and the consumption of processed food: A pragmatic randomised controlled trial in community-based adults

**DOI:** 10.1101/2021.01.29.21250725

**Authors:** Nicholas Edward Phillips, Julie Mareschal, Nathalie Schwab, Emily N.C. Manoogian, Sylvie Borloz, Giada Ostinelli, Aude Gauthier-Jaques, Sylvie Umwali, Elena Gonzalez Rodriguez, Daniel Aeberli, Didier Hans, Satchidananda Panda, Nicolas Rodondi, Felix Naef, Tinh-Hai Collet

## Abstract

Weight loss is key to control the increasing prevalence of metabolic syndrome (MS) and its components, i.e. central obesity, hypertension, prediabetes, and dyslipidaemia. We characterised the relationships between eating duration, unprocessed and processed food consumption, and metabolic health. During 4 weeks of observation, 213 adults used a smartphone application to record food and drink consumption, which was annotated for food processing levels following the NOVA classification. Consumption of unprocessed food items showed the highest number of significant associations with MS components after age and sex. In a pragmatic randomised controlled trial, we compared the metabolic benefits of 12h time-restricted eating (TRE) to standard dietary advice (SDA) in 54 adults with an eating duration >14h and at least one MS component. After 6 months, those randomised to TRE lost 1.6% of initial body weight (SD 2.9, p=0.01), compared to the absence of weight loss with SDA (−1.1%, SD 3.5, p=0.19). There was no significant difference in weight loss between TRE and SDA (between-group difference −0.88%, 95% confidence interval −3.1 to 1.3, p=0.43). Our results show the potential of smartphone records to predict metabolic health and highlight that further research is needed to understand individual responses to TRE.

## 1. Introduction

The prevalence of the metabolic syndrome (MS) is increasing worldwide, affecting approximately 1 billion people [1]. MS is a cluster of different clinical conditions (referred to as MS components, i.e. central obesity, elevated blood pressure, impaired glucose tolerance, and dyslipidaemia) which share common mechanisms [2]. This puts a substantial proportion of the population at risk for cardio-metabolic diseases, such as myocardial infarction, stroke, diabetes and their long-term complications [3].

Weight loss is a key factor to control MS components. Even a 5% weight loss is associated with metabolic benefits [4], but most patients struggle to achieve and maintain weight loss by lifestyle modifications (i.e. a healthier diet and increased physical activity). Current pharmacological options are not always efficacious and tolerated [5], and bariatric surgery is reserved for patients with severe obesity and requires close follow-up [4]. This explains the strong interest in new approaches to address isolated obesity (i.e. without comorbidities) or multiple MS components.

Recent animal and human studies have highlighted the role of *eating timing* in addition to the amount of *calorie intake*. These approaches are often called intermittent or periodic fasting and recommend various dietary regimens on daily, weekly or monthly timescales [6,7]. One form of intermittent fasting, called time-restricted eating (TRE), limits energy intake to certain daily time intervals without restrictions on calories or macronutrients [8,9]. In human studies, TRE can lead to weight loss even without deliberate calorie restriction [10–14], which could result from a loss of both lean and fat mass [15]. Changes to glucose metabolism have also been observed, with mixed effects on HbA1c [11,16], fasting glucose [17], insulin sensitivity [18], or the area under the curve of continuous glucose measurement [19]. An important and unaddressed question is whether TRE can lead to health benefits beyond the standard dietary advice (SDA) which is part of the clinical standard of care to address MS and isolated obesity [4].

Mobile technologies hold great promise to track daily eating behaviour and improve metabolic health as part of daily routine. They offer potential advantages over more traditional methods, such as food diaries or food questionnaires, by avoiding memory recall bias and minimising negative feedback and the Hawthorne observer effect [20]. In a landmark study using the smartphone application MyCircadianClock [10], Gill and Panda remotely collected data on eating behaviour to identify adults that may benefit from TRE. Once data are remotely collected, a key question is how to optimally characterise the relationship between the ingestion events and metabolic health. One possibility is to use photographs and annotations to estimate the number of calories in each consumed meal and drinks. This requires either asking participants to record exact weights and ingredients of meals [21], or a semi-quantitative estimation of all pictures by trained dietitians based on reference tables and standard recipes. Beyond calories and macronutrient content, food quality can be assessed using systems such as the NOVA classification, which uses four categories to rank food from unprocessed to ultra-processed [22–24]. To date, it is not established whether NOVA classification of food and drink events recorded by smartphone carries predictive information across multiple metabolic health indicators.

The goals of our study were two-fold. First, we characterised the relationships between eating duration, unprocessed and processed food consumption, lifestyle factors and metabolic health using a smartphone application in the Swiss adult population over 4 weeks. Second, we investigated whether a 12-hour TRE leads to metabolic benefits compared to SDA in a pragmatic randomised controlled trial lasting 6 months.

## 2. Materials and Methods

The SwissChronoFood trial is an open-label pragmatic randomised controlled trial evaluating the metabolic effects of TRE vs. SDA (clinicaltrials.gov no NCT03241121, Kofam.ch no SNCTP000002259). People aged ≥ 12 years were eligible, although only adults (≥ 18 years old) are reported in this article. Each participant signed a written consent form (see Institutional Review Board Statement below).

### 2.1. Study design and intervention

The study comprised an *observation phase* to assess the baseline metabolic parameters and the daily eating behaviour of community-based adults using a research-dedicated smartphone application over 4 weeks. We recruited via posters, online adverts and social media in two different languages and cultural regions of Switzerland, i.e. at Lausanne University Hospital, Lausanne and at Inselspital, Bern. Inclusion criteria were adults with a body mass index (BMI) ≥ 20 kg/m^2^, stable weight (± 2 kg) over the previous 3 months, and the regular and confident use of a smartphone compatible with the MyCircadianClock application (iOS or Android systems) [10].

After 4 weeks of observation, individuals with an eating duration (defined below) of > 14 hours and had at least one component of MS were eligible for the intervention phase (**Figure S1**). The criteria of each MS component followed the International Diabetes Foundation (IDF) consensus definition [2]: Central obesity was defined as BMI ≥ 30 kg/m^2^ or waist circumference (WC) ≥ 80 cm (women) or WC ≥ 94 cm (men); hypertension as systolic blood pressure (BP) ≥ 130 mmHg and/or diastolic BP ≥ 85 mmHg; impaired fasting glucose for plasma levels ≥ 5.6 mmol/L (100 mg/dL); high triglycerides for fasting plasma levels ≥ 1.7 mmol/L (150 mg/dL); and low HDL cholesterol for fasting plasma levels < 1.29 mmol/L (50 mg/dL, women) or < 1.03 mmol/L (40 mg/dL, men).

In the *intervention phase*, participants were randomised to the TRE or SDA intervention, with a 1:1 allocation ratio and block randomisation of 8 units stratified by sex and recruitment site. The intervention could not be blinded, but the allocation table was computer-generated and concealed. Those in the TRE arm were asked to limit their consumption of food and calorie-containing drinks to a 12-hour window of their choice, with no advice on nutrition quality, quantity of macronutrients or calorie intake. Those in the SDA arm received brief nutritional counselling and were given a leaflet summarising the food pyramid and Swiss recommendations for a balanced and healthy diet [25,26], which is considered the standard of care. After 6 months, the metabolic measurements were repeated at the closeout visit. In addition, participants were contacted at 2 and 4 months post-randomisation to self-report their body weight and address compliance with the intervention.

In addition to the eligibility criteria of each study phase listed above, we excluded pregnant and breastfeeding women, individuals with a major illness/surgery over the previous month, active cancer or under treatment over the previous 12 months, eating disorder, prior bariatric surgery, those who had been following a weight management programme over the previous 3 months, shift workers, and those travelling to a different time zone during the observation phase. Additionally for the intervention phase, we excluded adults with cardiovascular disease (e.g. coronary heart disease, cerebrovascular disease, peripheral artery disease), major liver, gastrointestinal, renal, endocrine or sleep disorders, diabetes mellitus with hypoglycaemic drug(s), those on centrally acting medications (benzodiazepines, zolpidem, zopiclone, antidepressants, neuroleptics and psychotropic drugs), lipid-lowering drugs, and medications affecting gut absorption, transit or weight.

#### Modifications to the study protocol

In the original protocol, we planned to randomise only participants with MS following the IDF consensus definition, i.e. those with central obesity *and* at least two other MS components [2]. This population proved difficult to recruit because the Swiss adult population has a lower prevalence of obesity than in most Western countries [27], thus leading to an even lower prevalence of MS. The inclusion criteria for the intervention phase were relaxed after 5 months of recruitment: those with an eating duration > 14 hours and had at least one component of the metabolic syndrome (MS) were eligible for the intervention phase (**Figure S1**).

### 2.2. Recording food and drink events with a smartphone application

Participants were instructed to take pictures of all consumed food, drinks and medications with the research-dedicated myCircadianClock smartphone application [10]. Recorded entries included a timestamped picture and a free-text annotation. Participants could annotate photographs either immediately or in the following hours. Optionally, participants could type text-only entries without any picture, e.g. if the smartphone ran out of battery, or if it was not socially acceptable to take pictures in the current context.

The *eating duration* was calculated from the timestamp of all recorded ingestion events (food items, drinks, except for water and medications) during the observation phase [10,15]. In order to be less sensitive to outliers (special days with time shifts in ingestion events) or entry errors, the eating duration was defined as the time interval between the 2.5^th^ and the 97.5^th^ percentiles of all timestamped ingestion events. To account for the social consumption habits and the nadir of food entries, we calculated the percentiles using a start time of 04:00a.m. as defined previously [10], which also corresponded to a low point of ingestion events in this study (**Figure S2**). The eating duration of the intervention phase was estimated in a similar fashion. Participants did not receive feedback on their eating duration until the end of the observation phase. They were regularly reminded to record ingestion events at planned regular phone calls or via email after 2 weeks of observation, and after 2 and 4 months of intervention (**Figure S1**).

### 2.3. Categorising ingestion events based on free-text annotations

We developed an analysis pipeline using Python scripts to categorise the free-text annotations of recorded ingestion events in the observation and intervention phases. To ensure good coverage and improve the number of text entries with an assigned NOVA category, we performed the following pre-processing steps: 1) Each text entry containing multiple food items separated by commas were split into multiple text entries; 2) Numbers, semi-quantitative terms (e.g. “slice,” “glass,” “peace,” “half”), units (e.g. “grams,” “decilitres”) and multiple blank spaces were removed. 3) We then counted the number of unique text entries using the *Tokenizer* function in Keras (Python module, version 2.2.4). Free-text annotations sometimes comprised typos and spelling variations, and to correct for this we further processed words based on the Levenshtein distance (Python module python-Levenshtein, version 0.12.0). The Levenshtein distance between two words is the minimum number of single-character changes (substitutions, insertions or deletions) that is required to transform one word into the other. For any two entries with a Levenshtein distance of one, the entry with the largest number of counts was preferred e.g. “ham piza” would be transformed into “ham pizza” if the second entry had more counts across all recordings. 4) Finally, text entries that were recorded only once were not included in this analysis. From a total of 17,438 unique entries, 11,030 were recorded once and 6408 were recorded more than once, but this filtering still gave high coverage due to the fact that some items are frequently used. For example, the top entry “cafe” (“coffee” in English, “Kaffee” in German, and many other spelling variants) was used 3031 times.

We assessed food processing according to the qualitative NOVA classification, which uses four categories to rank food from unprocessed to ultra-processed [22–24]. No quantitative assessment of the macronutrient intake, number of calories, or types of food items was attempted. In addition to the 4 NOVA categories (NOVA1: “Unprocessed or minimally processed foods,” NOVA2: “Processed culinary ingredients,” NOVA3: “Processed foods,” NOVA4: “Ultra-processed foods”), we added categories for beverages grouped into “Alcohol-containing drinks” (A), “Caffeinated drinks” (C), “Sweet drinks” (S), and “Other drinks” (D). With this extended classification, each drink entry could therefore be assigned multiple categories. For example, the soda Coca-Cola was an ultra-processed, caffeinated and sweet drink (abbreviated NOVA4-CS), coffee with sugar and milk was a NOVA2, caffeinated and sweet drink (abbreviated NOVA2-CS), and a black tea was a NOVA1, caffeinated drink (abbreviated NOVA1-C).

The pre-processed text entries were annotated using the extended NOVA classification by 4 independent reviewers (J.M., N.E.P., S.U., T.H.C.). A category was assigned to a given entry if a minimum of 3 reviewers selected the category. A consensus was reached for the remaining entries by at least 3 reviewers. For entries collected in German (at the Bern research site), representing 16.4% of all entries, the entries were annotated by a single reviewer (T.H.C.) due to language considerations. Some food entries required assumptions on their base recipes and ingredients. Items were assumed to be homemade unless stated otherwise [28,29], with limited exceptions (e.g. chocolate-containing food and drinks, biscuits, toast and soft bread, croissants, pizza, burgers, plant-based drinks) because they most often involve food processing. A single dish composed of multiple items and ingredients was assigned the highest single NOVA category based on their base recipe.

### 2.4. Quantifying compliance to the study intervention

For participants randomised to TRE or SDA, we quantified their compliance to the allocated intervention by comparing their ingestion events during the intervention phase to those recorded during the observation phase. We quantified the eating duration (defined in section 2.2) and the percentage of ingestion events belonging to each NOVA and drink category (defined in section 2.3). This was done separately for the observation and intervention phases to calculate the changes (Δ) pre- and post-randomisation. The compliance of participants in the TRE arm was assessed according to their eating duration in the intervention phase. Participants in the SDA arm were considered compliant if the proportion of ingestion events with NOVA4 category decreased and the proportion of ingestion events with NOVA1 categories increased.

### 2.5. Primary and secondary outcomes, other measurements

The primary outcome of the study was the change in weight after 6 months following the 12-hour TRE vs. SDA intervention. Weight was measured in light clothing with calibrated medical-grade scales at baseline (visit 1), at randomisation (visit 3) and after 6 months of follow-up (visit 6, **Figure S1**). In addition, weight was measured at home and recorded at interim phone calls (visits 4 and 5). Because these weights were self-reported, they were analysed in a secondary analysis.

The secondary outcomes were the changes in body mass index (BMI), WC, BP, and fasting plasma concentrations of glucose, HDL cholesterol and triglycerides. The BMI was calculated as the weight in kilograms divided by the height in metres squared. The height was measured barefoot using a calibrated stadiometer. BP was measured three times with a calibrated BP monitor (Omron device) and an appropriately sized arm cuff, after 5 minutes of rest in the sitting position, and the last two values were averaged.

We measured other metabolic parameters, such as glycated haemoglobin (HbA1c), using standard biochemistry assays at the same time points. Body composition was measured by whole-body dual energy X-ray absorptiometry (DXA, GE Healthcare Lunar iDXA at Lausanne site, GE Healthcare Lunar Prodigy Advance at Bern site) following international guidelines [30]. With minimal ionising radiation, DXA can assess total lean mass, fat mass, and body fat percentage [31]. The visceral adipose tissue (VAT) was computed by subtracting subcutaneous adipose tissue from the total android fat mass [32]. At baseline, sleep quality was assessed with the Pittsburgh Sleep Quality Index (PSQI) [33] and physical activity with the International Physical Activity Questionnaire (IPAQ) [34]. Sleep duration and the midpoint of sleep were assessed on work days and free days using the Munich Chronotype Questionnaire [35].

### 2.6. Statistical analyses

Data are reported as mean ± standard deviation (SD), or median (interquartile range, IQR) if not normally distributed. Comparisons between groups or sex were calculated with the Chi^2^ statistic (or exact Fisher test where appropriate) for categorical variables, or Student t-test for continuous variables. Paired sample t-tests were performed (Scipy module in Python, version 1.2.1; Stata software package, version 16.1) to test for the null hypothesis that pre- and post-intervention measurements had identical mean values. Non-paired t-tests were used to compare the differences between the TRE and control group (which have different numbers of participants). A two-tailed p-value <0.05 was considered statistically significant. All analyses were performed with the intention-to-treat approach, although we also assessed weight changes in a secondary per-protocol analysis.

Sample size calculations were based on general assumptions as no randomised controlled trial of TRE was published at the time of study design. Gill and Panda reported a weight loss of 3.27 kg (approx. SD 1.18) after TRE [10] and we expected the standard of care SDA to result in a weight loss of 1.0 kg (approx. SD 1.0), although this varies depending on the participant’s motivation and the intervention intensity. Using an alpha level of 0.05, a power of 90% and an estimated attrition rate of 25%, this difference was expected to be observed with 20 participants (10 in the TRE arm, 10 in the SDA arm).

Bayesian linear regression [36] was performed using data from the observation phase to quantify the relationship between the explanatory variables and the outcome variables (clinical measurements related to metabolic health). The explanatory variables included age, sex, the number of ingestion events for each of the NOVA and drink categories, eating duration, the midpoint of sleep on work/free days, sleep duration on work/free days, sleep quality (PSQI), and physical activity (IPAQ). For each of the participants *i =* 1,…, *N* there is a set of *K* explanatory variables *x*_*i*_ *=* (*x*_*i*1_,…,*x*_*iK*_) and a set of, *D* clinical measurements *y*_*i*_ *=* (*y*_*i*1_,…, *y*_*iD*_). The linear regression model for each clinical measurement *d* for participant *i* is then given by

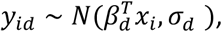

where for each clinical measurement *d* there is a vector of regression coefficients *β*_*d*_ *=* (*β*_*d*1_,…, *β*_*dK*_) and a parameter *σ*_*d*_ to quantify the SD of the observation error. The clinical measurements were log-transformed before further analyses to make the distributions closer to a normal distribution, and the IPAQ questionnaire result was transformed using a square root operation. We normalised both the clinical measurements and the explanatory variables by subtracting the mean and dividing by the standard deviation. In the Bayesian regression, we used priors of *σ*_*d*_ ∼ *N*(0,1) and *β*_*dk*_ ∼ *N*(0,0.1) to regularise the inference problem. Posteriors for the regression coefficients and parameters were inferred by the Hamiltonian Monte Carlo sampler in the STAN probabilistic programming language [37] with 4 different chains of 1000 samples each. To perform variable selection, we fitted the model with all variables and then only selected variables if 95% of the parameter estimates were greater or less than one. To validate predictive performance of the regression models we performed leave-one-out cross-validation (LOO-CV) whereby a test participant was removed from the data and the model was optimised using the remaining participants. The *R*^2^ value was then calculated using the predictions on the test participants.

For participants randomised to TRE or SDA, we performed a secondary analysis using the weight measurements at visits 1 and 3 (the observation phase) and visit 6 (6 months after the start of intervention), as well as the self-reported weights at visits 4 and 5 (2 and 4 months after start of intervention, respectively). For each participant, we could then estimate the change in weight using the following quadratic model

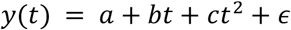

where *y*(*t*) is the weight measurement (kg) at time *t, t* is time in months after the start of intervention, *a* is the starting weight (kg), *b* is the regression coefficient for the linear rate of weight loss (in kg/per month) while *c* is the regression coefficient for the quadratic term (in kg/per month^2^), and *ϵ* is a normally distributed noise term with mean 0 and standard deviation *σ*_*ϵ*_ to account for measurement noise and natural fluctuations in weight. The quadratic term was included to account for weight time series that showed a nonlinear, curved profile. The two weight measurements in the observation phase (visits 1 and 3) are considered as *t =* 0. We used weakly informative priors on the parameters *b, c, σ*_*ϵ*)_ ∼ *N*(0,10) and again used sampling within STAN to estimate the parameters. After performing parameter inference, this approach provides confidence bounds on the weight change for each participant.

## 3. Results

### 3.1. Recruitment of participants

Out of the 729 adults screened for eligibility, 511 did not meet the criteria (detailed in **Figure 1**). We included 218 adults in the observation phase, but had to exclude 5 individuals just after inclusion for undisclosed psychiatric disorders, or a BMI lower than self-reported. Accounting for these exclusions, the observational data of 213 adults could be analysed. Next, we randomised 28 participants to TRE and 26 to SDA intervention. Two individuals allocated to the SDA arm had to be excluded the day after randomisation [38], when it was learnt that they met an exclusion criteria for the intervention phase (regular use of statin or antidepressant).

**Figure 1.**
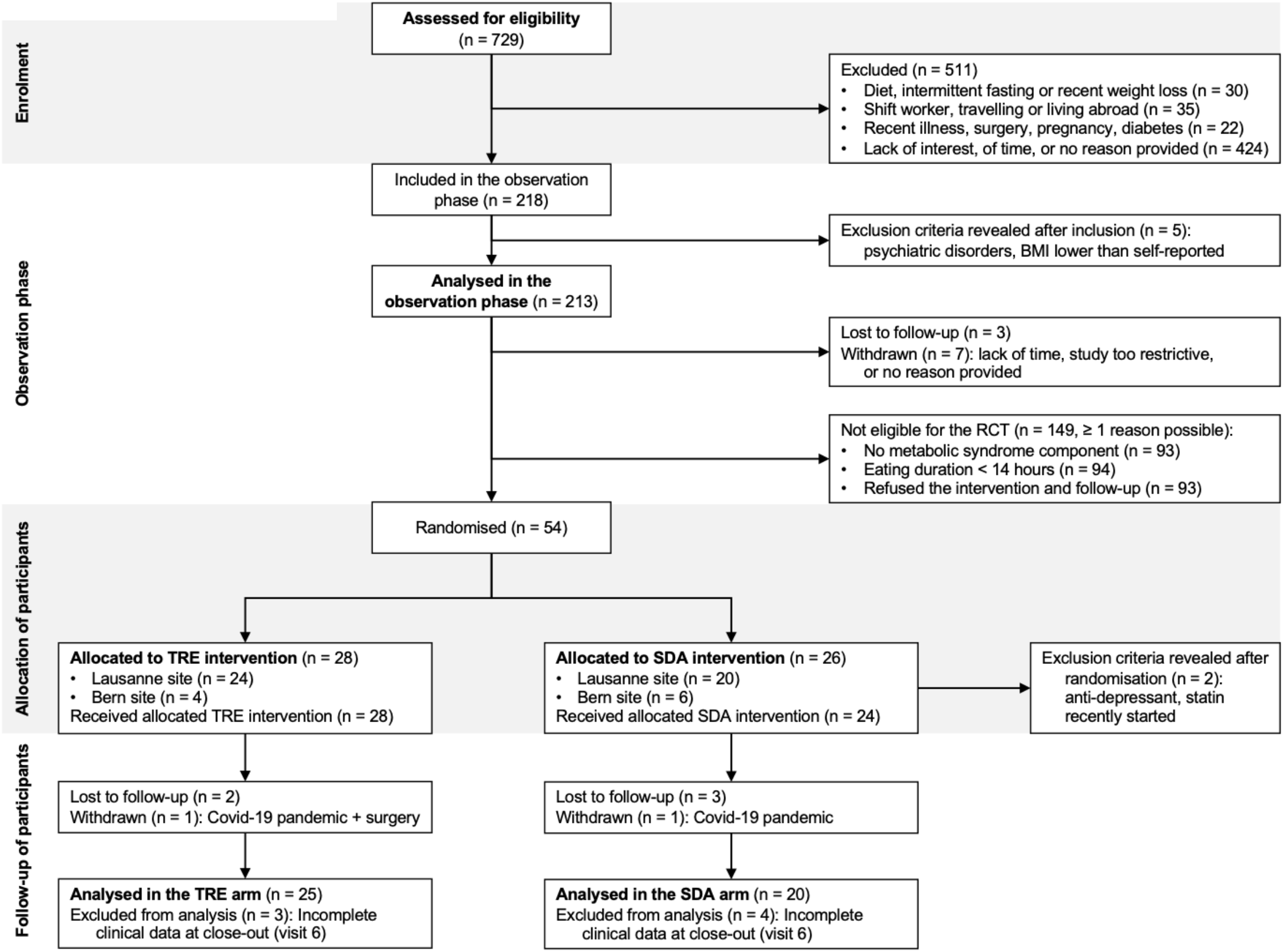
Flow of participants. <u>Legend:</u> Adapted from [44]. Out of the 729 adults screened for eligibility, 213 were analysed in the observation phase and 54 were randomised to 12-hour time-restricted eating (TRE) vs. standard dietary advice (SDA). During the intervention phase, 2 participants had to be excluded the day after randomisation (see text), 5 were lost to follow-up and 2 withdrew from the study (due to Covid-19 pandemic or other reasons). The main analyses therefore comprised 25 participants in the TRE arm and 20 participants in the SDA arm.

### 3.2. Baseline characteristics of the participants

The 213 adults in the observation phase had a mean age of 40.1 years (SD 13.3, range from 18 to 77) and 71% were women (**Table 1**). The median BMI was 24.9 kg/m^2^ (IQR 22.6–29.1). Central obesity was found more often in women (55%) than in men (39%, p=0.04). On the other hand, hypertension, impaired fasting glucose and elevated triglycerides were more prevalent among men than women (all p≤0.002). While the mean fasting plasma glucose was 5.2 mmol/L (SD 0.6) in men and 4.9 mmol/L (SD 0.5) in women (p=0.0001), the HbA1c levels were comparable between sex (p=0.19).

**Table 1.**
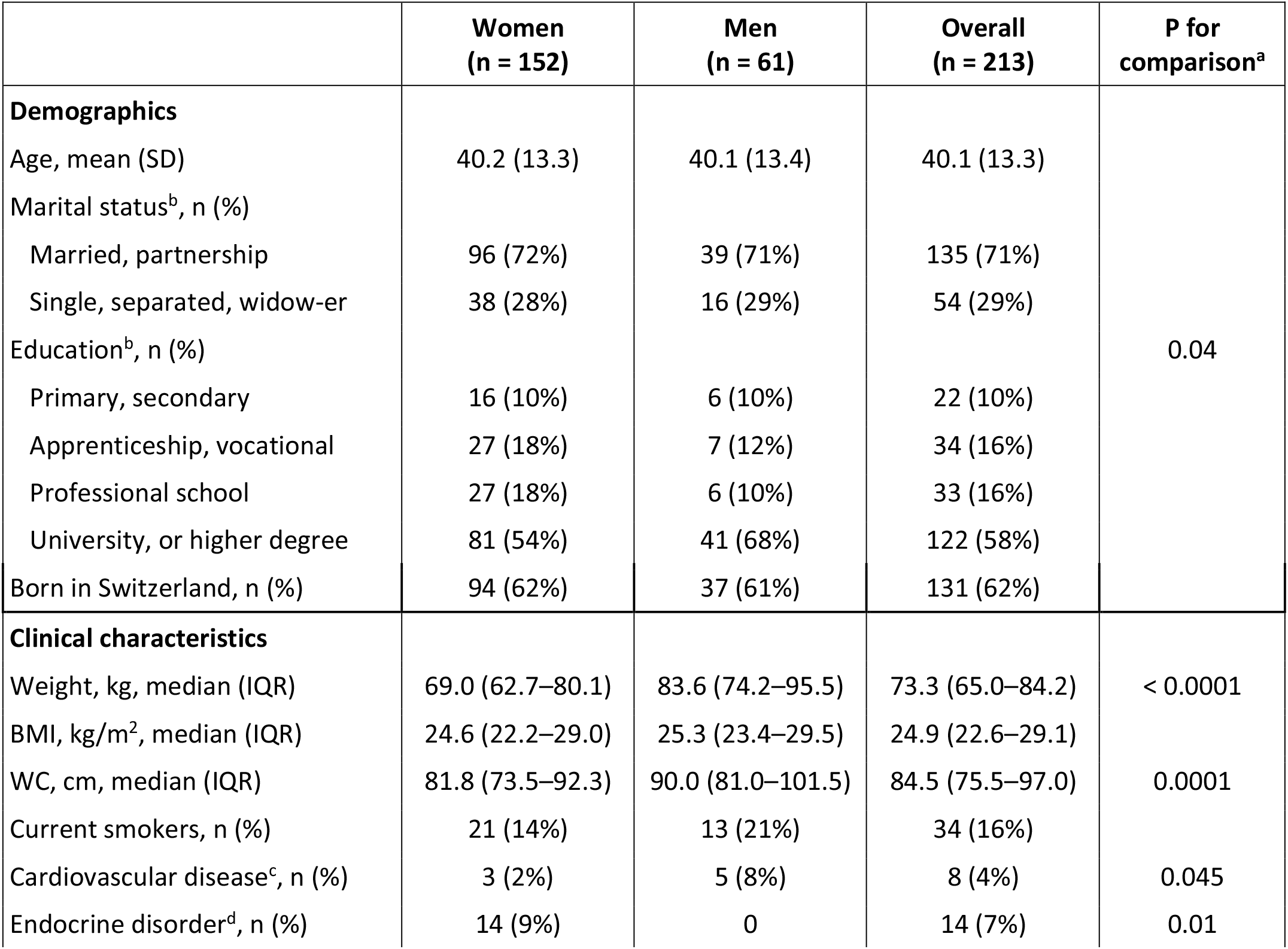
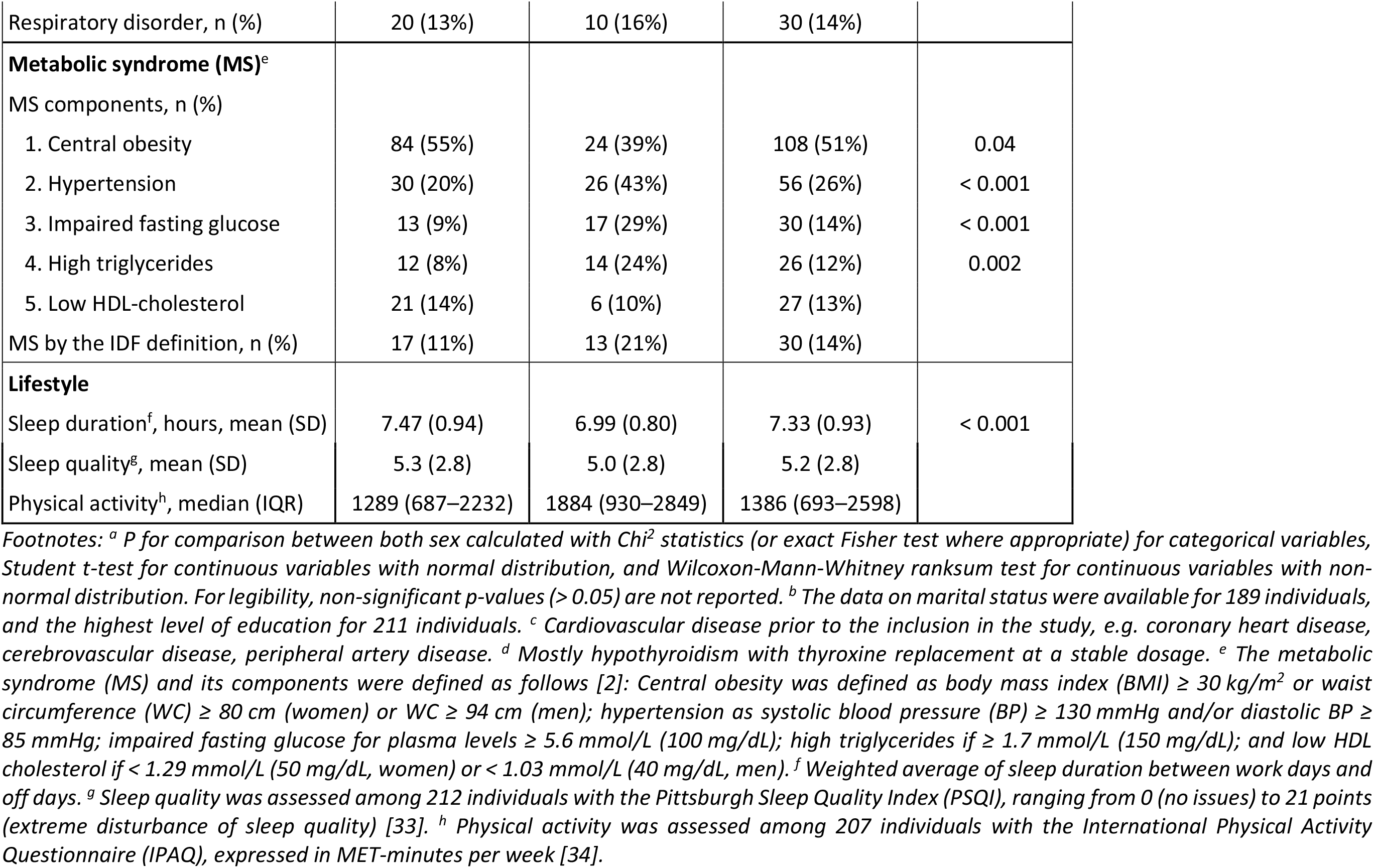
Baseline characteristics.

Following the IDF definition [2], the prevalence of MS was not statistically different between men (21%) and women (11%, p=0.055). When looking at MS components separately, 38% had no MS component, 27% a single MS component, 21% two MS components, and 14% three or more MS components. We therefore decided to include participants with at least one MS component at baseline (section 2.1).

### 3.3. Relationship between metabolic health, unprocessed and processed food consumption at baseline

A total of 88.7% of ingestion events (out of 102,072 events) were annotated according to the expanded NOVA classification, of which 2.4% were removed as they were water or flavoured water, 1.2% were medications, and 1.5% had text that was not sufficiently informative to be fully annotated. The remaining 11.3% of ingestion events were not annotated because they represented single entries. Of the annotated ingestion events, unprocessed or minimally processed foods (NOVA1) represented 45.9%, processed culinary ingredients (NOVA2) 13.1%, processed foods (NOVA3) 18.2%, and ultra-processed foods (NOVA4) 22.9%. Drink entries were also classified according to the newly devised categories: Alcohol-containing drinks (NOVA-A) represented 2.9% of the annotated entries, caffeinated drinks (NOVA-C) 11.1%, sweet drinks (NOVA-S) 2.5%, and other drinks (NOVA-D) 5.0%. Some entries were designated multiple categories, and the overlap between different categories is shown in **Table S1**.

We next examined whether age, sex, eating duration, NOVA classification of ingestion events, and questionnaires on sleep and physical activity contained predictive information for each metabolic health parameter at baseline in a Bayesian linear regression. We only report here the significant coefficients (**Figure 2A**). The regression coefficient for age was significant for all metabolic health parameters except HDL cholesterol. The regression coefficient for sex was significant for 5 out of 8 of the metabolic health parameters, and the estimated coefficients for women were negative for WC, systolic BP, triglycerides and glucose, which is in line with their lower values typically seen among women. Out of the ingestion events, the number of unprocessed food items (NOVA1) had the highest predictive performance. It was negatively associated with BMI and diastolic BP, and positively associated with HDL cholesterol. The ingestion of ultra-processed food items (NOVA4) was positively associated with WC. The consumption of alcohol-containing drinks (NOVA-A) was positively associated with HDL cholesterol, and both caffeinated (NOVA-C) and sweet drinks (NOVA-S) with triglyceride levels. From the sleep questionnaires, a negative relationship was found between sleep duration on work days and BMI and WC. Finally, the level of physical activity (IPAQ) was negatively associated with BMI, WC and triglycerides, and positively associated with HDL cholesterol.

**Figure 2.**
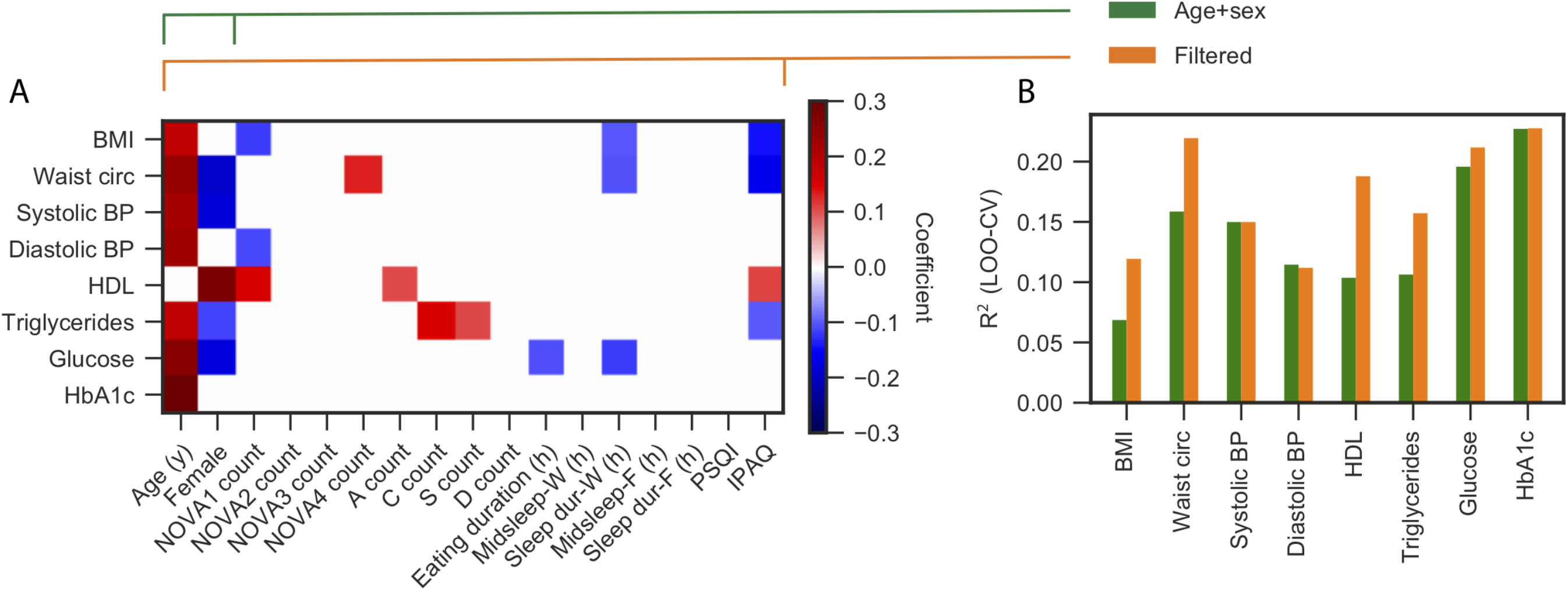
The addition of NOVA categories and physical activity to age and sex increases the predictive power for MS components. <u>Legend:</u> (A) Each row shows the regression coefficients for predicting the clinical variable (labelled on the y-axis) using the explanatory variables (labelled on the x-axis), all measured in the observation phase. BMI, body mass index; BP, blood pressure; HbA1c, glycated hemoglobin; HDL, high-density lipoprotein cholesterol; NOVA count, the number of ingestion events of each NOVA category; NOVA-A, alcohol-containing drinks; NOVA-C, caffeinated drinks; NOVA-S, sweet drinks; NOVA-D, other drinks; Midsleep-W and Midsleep-F, the midpoint of sleep on work days and free days, resp.; Sleep dur-W and Sleep dur-F, the sleep duration on work days and free days, resp.; PSQI, Pittsburgh Sleep Quality Index [33]; IPAQ, International Physical Activity Questionnaire [34]. The regression coefficients show the mean posterior parameter estimate, and variables that are not significant are shown as white space. Red colour corresponds to a positive coefficient, blue colour a negative coefficient. (B) Bar heights represent the R^2^ values calculated on test participants using leave-one-out cross-validation (LOO-CV) to compare the predictive performance of a model that uses only age and sex (green bars) vs. a model that uses all variables that were significant in panel A (orange bars, selected variables shown in panel A).

Given that age and sex were frequently found to have significant regression coefficients (**Figure 2A**), we wanted to confirm that the inclusion of the additional explanatory variables from ingestion events and questionnaires increased predictive performance. Indeed, the R^2^ values calculated on test participants using leave-one-out cross-validation (LOO-CV) showed that inclusion of those variables increased predictive performance for BMI, WC, HDL cholesterol and triglycerides, while there was no improvement in performance over age and sex alone for systolic and diastolic BP, glucose and HbA1c (**Figure 2B**). Of note, there was a negative relationship between eating duration and sleep on work days and glucose levels, but this did not lead to substantially increased predictive performance when these variables were included.

### 3.4. Compliance with the randomly assigned interventions

We evaluated the changes in the fraction of NOVA categories and in eating duration between study phases to quantify compliance to each intervention (TRE and SDA) after randomisation.

Relative to baseline, those in the SDA arm significantly increased the proportion of unprocessed or minimally processed food (NOVA1, +7.0%, SD 9.7, p<0.01) and significantly decreased the proportion of ultra-processed food (NOVA4, −7.6%, SD 11.0, p<0.01, **Figure 3A**). The increased consumption of NOVA1 items and decreased consumption of NOVA4 items in the SDA group were significant compared to TRE (p<0.01 and p=0.04, respectively, **Table S2**). No significant changes were observed for the consumed drink categories.

**Figure 3.**
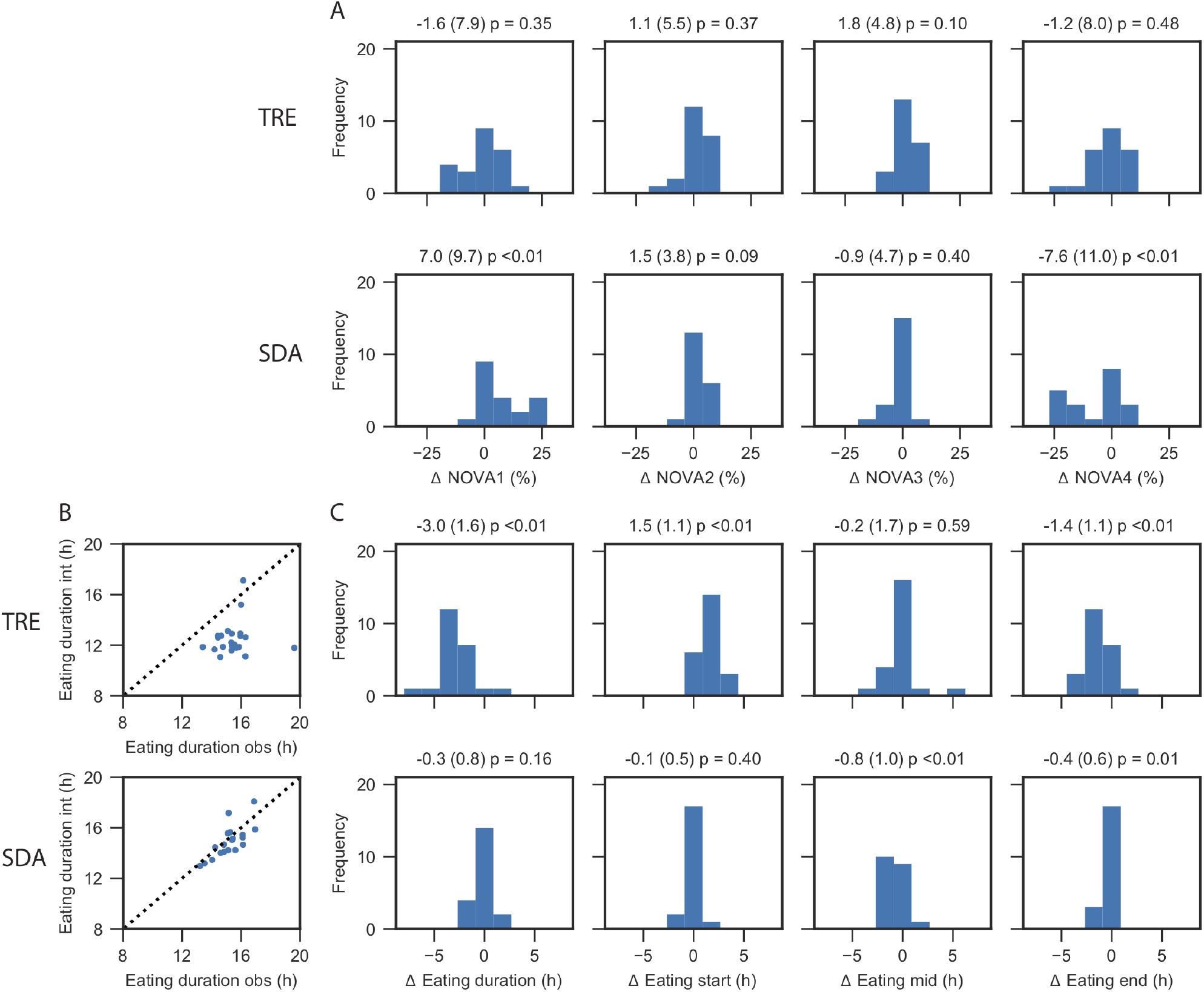
Changes in the consumption of NOVA categories and in eating duration with the TRE and SDA interventions. <u>Legend:</u> During the 6-month intervention phase, the changes in consumption of NOVA categories (panel A) and in eating duration (panels B-C) showed compliance with time-restricted eating (TRE) and standard dietary advice (SDA) interventions. Numbers above each histogram represent the changes (unit on the x-axis) as mean, standard deviation and p-value. Panel A, from left to right: Changes (x-axis) in the proportion of unprocessed or minimally processed foods (NOVA1), processed culinary ingredients (NOVA2), processed foods (NOVA3), and ultra-processed foods (NOVA4), depicted as % of all classified ingestion events and the number of participants per bin is shown on the y-axis. Panel B: Scatter plot of eating duration during the observation and the intervention phases with TRE (top) and SDA (bottom). Panel C, from left to right: Changes (x-axis) in eating duration, the start of the eating interval, the median of the eating interval, and the end of the eating interval compared to the observation phase, in fractional hours (x-axis, i.e. 1.33h = 1 hour and 20 minutes). A shift to the left means a shorter eating duration, an earlier start, an earlier median, and an earlier end of the eating interval, resp., while a shift to the right means a longer eating duration, a later start, a later median, and a later end of the eating interval, resp.

The timestamp of each ingestion event allowed the assessment of changes in eating duration during the intervention phase (**Figure 3B-C**). The TRE intervention shortened the eating duration compared to baseline (−3.0 h, SD 1.6), which was due to a later start of the eating interval (+1.5 h, SD 1.1) and an earlier end of the eating interval (−1.4 h, SD 1.1, all p<0.01). Changes of the eating interval after TRE were all statistically significant compared to SDA (all p<0.01 for pairwise comparisons, **Table S2**).

The changes in NOVA categories in the SDA arm on the one hand, and changes in eating duration in the TRE arm on the other hand, confirm that participants followed the randomly allocated intervention until the closeout visit.

### 3.5. The effects of TRE vs. SDA on clinical outcomes

We measured the changes in clinical outcomes relative to baseline (**Table S3**) and compared the metabolic effects of TRE and SDA. After 6 months, those randomised to TRE lost 1.6% of initial body weight (SD 2.9, p=0.01), compared to the absence of weight loss with SDA (−1.1%, SD 3.5, p=0.19, **Figure 4A**). Thus, there was no significant difference in weight loss between TRE and SDA (between-group difference −0.88%, 95% confidence interval −3.1 to 1.3, p=0.43, similar results with non-parametric tests). Similar results were obtained in the per-protocol analysis (between-group difference −1.93%, 95% confidence interval −4.0 to 0.1, p=0.09).

**Figure 4.**
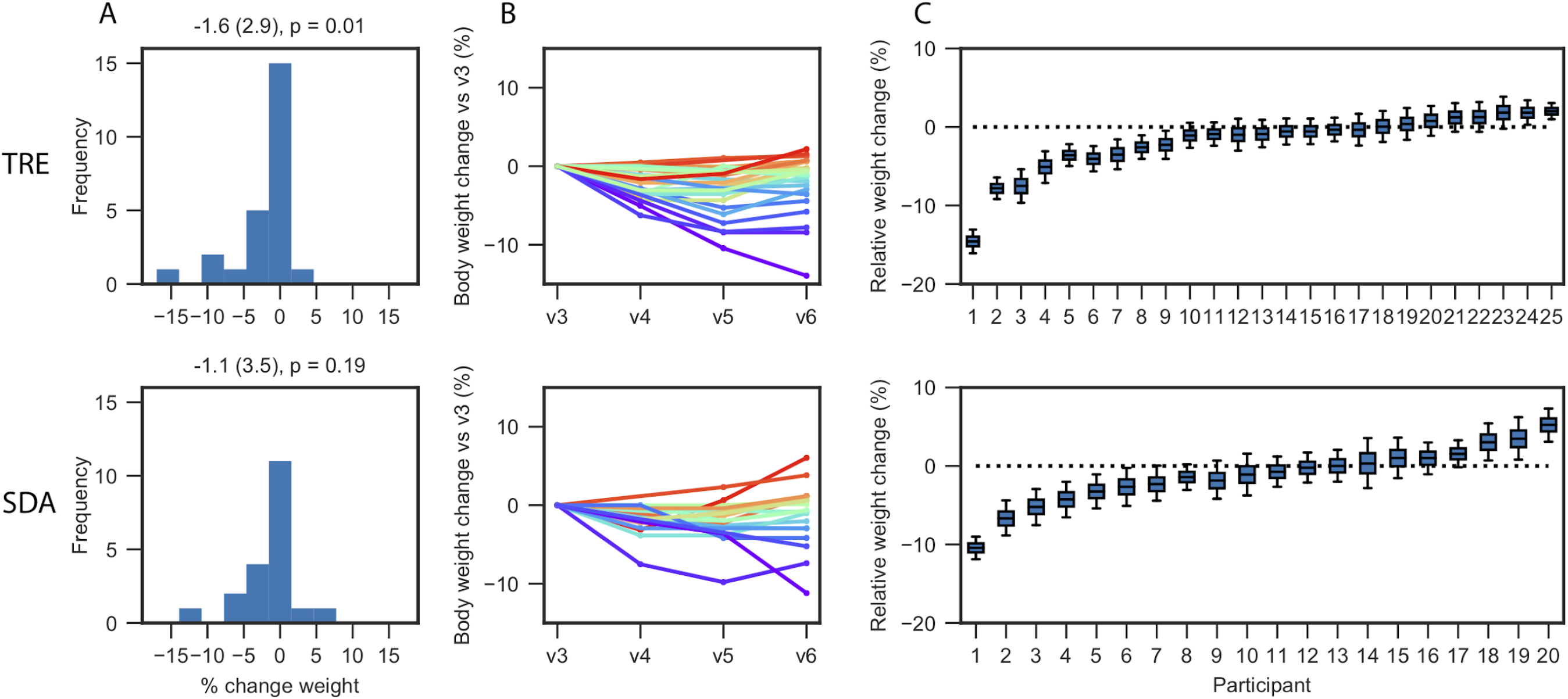
TRE alters body weight. <u>Legend:</u> (A) Change of body weight after time-restricted eating (TRE, top) compared to standard dietary advice (SDA, bottom). Data are presented as histograms of weight change (in % of initial body weight). Numbers above the histograms represent the weight change as mean, standard deviation and p-value. (B) Recorded weight measurements for each participant across multiple visits expressed as a percentage change relative to initial body weight. (C) For each individual, the relative weight change is estimated using all recorded weight measurements and a quadratic regression model. The whiskers show the 5^th^ and 95^th^ percentiles of estimated weight change, while the boxes show the 25, 50, and 75 percentiles.

While the average change in weight in both arms was modest, some participants lost close to 10% of initial body weight (**Figure 4B**). We next used all recorded weights to model the weight loss for each individual. We estimate that in the TRE group 2 participants significantly gained weight and 9 lost weight, whereas in the SDA group 3 participants significantly gained weight and 6 lost weight (**Figure 4C**).

We also assessed whether differences were observed in the secondary outcomes, i.e. changes in BMI, WC, systolic and diastolic BP, HDL cholesterol, triglycerides, glycated haemoglobin, fasting plasma glucose levels (**Figure S3**), and body composition (**Figure S4**). However, the observed changes in these metabolic outcomes were not statistically significant between TRE and SDA (**Table 2**). Finally, there were no major changes in the number of MS diagnoses following the full IDF definition. For those with complete data, in the TRE group 5 out of 22 participants had MS at baseline, and one more participant was diagnosed with MS after intervention. In the SDA group, 5 out of 18 participants had MS at baseline, and one participant “gained” MS diagnosis and one participant “lost” MS diagnosis after 6 months.

**Table 2.**
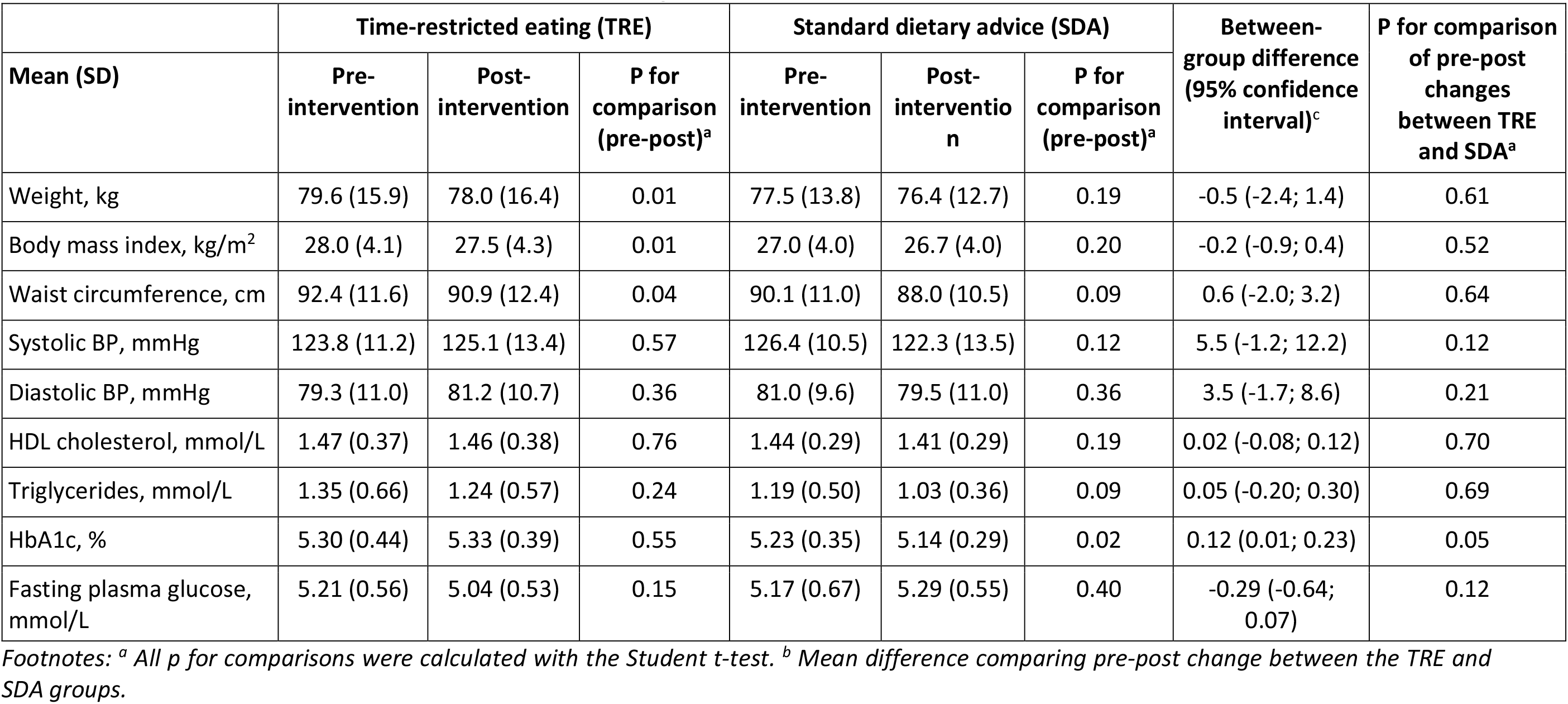
Effects of TRE and SDA on clinical outcomes and eating behaviour.

### 3.6. Weight loss during the TRE intervention is associated with the number of events recorded in the observation phase

Finally, we assessed potential explanatory variables that were associated with increased weight loss in the TRE group. While we found no significant association with either sex, age, weight at baseline, reduction in eating duration or the eating duration during the intervention phase (**Figure 5**), we found a positive correlation between the percentage of weight loss and the number of ingestion events recorded during the observation phase (p=0.01, **Figure 5G**).

**Figure 5.**
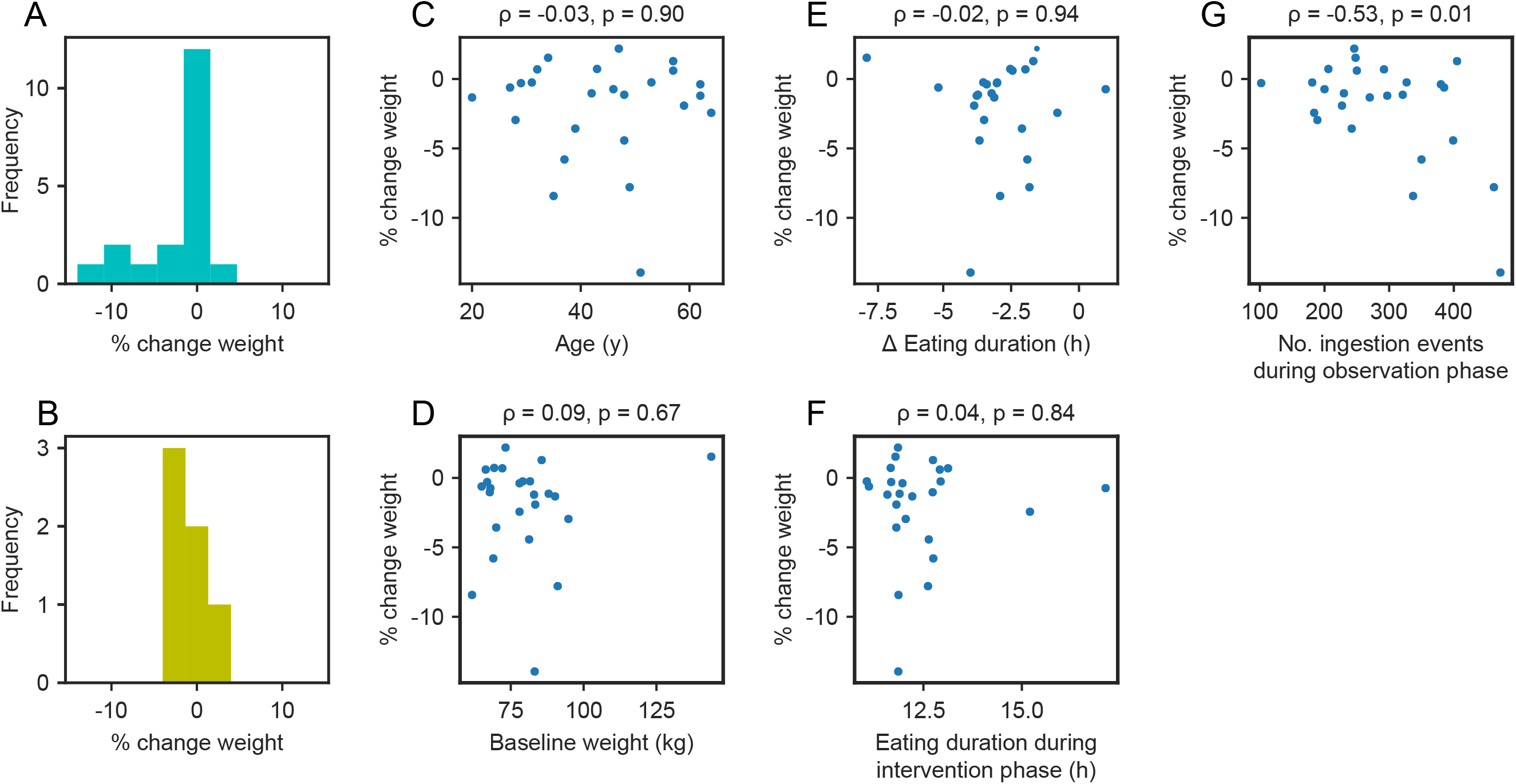
The number of items recorded in the observation phase correlates with weight loss. <u>Legend:</u> Exploration of potential explanatory parameters for more weight loss in the time-restricted eating (TRE) group: Subgroup analysis by sex (panel A: women; panel B: men, p for comparison=0.57), or weight change relative to age (panel C), weight at baseline (panel D), eating duration change between study phases (panel E), eating duration during the intervention phase (panel F), or the number of ingestion events recorded during the observation phase (panel G, after pre-processing of data as detailed in section 2.3). Numbers above the scatter plots represent the Pearson correlation coefficient (ρ) and the p-value.

## 4. Discussion

In our study, we characterised the relationship between eating duration, unprocessed and processed food consumption, and metabolic health. We found that the number of unprocessed food events was positively associated with HDL cholesterol and negatively associated with BMI and triglycerides. Using significant variables from the NOVA categories, sleep and activity questionnaires improved metabolic health modelling. Next, we investigated whether a randomised 6-month dietary intervention could lead to weight loss in a pragmatic community-based setting. Adults with at least one MS component and eating duration > 14 hours lost 1.6% (SD 2.9) of body weight with a self-selected 12-hour TRE, keeping in mind that this intervention group was not provided information about standard dietary guidelines. There was no significant difference in weight loss between TRE and SDA (between-group difference −0.88%, 95% confidence interval −3.1 to 1.3, p=0.43).

Mobile technologies hold the potential to track daily behaviour and improve metabolic health. This novel approach offers advantages over more traditional methods, such as food diaries or food questionnaires, as it avoids memory recall bias and minimises negative feedback and the Hawthorne observer effect [20]. Close monitoring of eating behaviour can only be achieved in closed facilities, but this limits the applicability of findings to real-life settings. Furthermore, smartphone applications can offer more flexibility to participants by allowing photos to be taken and then annotated at a later time, and the record of consumed meals can be hidden in order to minimise feedback that may lead to behavioural changes, which has been successfully used in previous studies [10]. In addition to observing community-based adults, the data from smartphone apps can help identify participants that may benefit from dietary interventions and subsequently evaluate compliance.

In our study, the ingestion events were recorded remotely with a smartphone application and their level of food processing was assessed with the NOVA classification. We found a positive correlation between ultra-processed food (NOVA4) consumption and waist circumference. This is in line with the association of ultra-processed food consumption and obesity in epidemiological studies in Sweden [39] and the US [40]. In a large longitudinal French cohort, higher consumption of ultra-processed food was associated with increased BMI and a higher risk of becoming overweight or developing obesity [41]. At the other end of the spectrum of food processing, the number of unprocessed food items (NOVA1) showed the highest number of significant relationships with MS components after age and sex in our study. The number of unprocessed food events (NOVA1) was positively associated with HDL and negatively associated with BMI and triglycerides. Our findings match the recommendations favouring unprocessed or minimally processed food in everyday diet and limiting processed food to curtail the obesity epidemic and improve metabolic health [41].

The NOVA classification does not specifically address drinks. Beverages are spread across NOVA categories following their level of processing: Coffee and tea are considered unprocessed (NOVA1), wine and beer processed through fermentation (NOVA3), while soda, energy drinks, and spirits are ultra-processed (NOVA4). We propose here an extension to the NOVA classification [22–24], by splitting all ingestion events with beverages into alcohol-containing drinks (NOVA-A), caffeinated drinks (NOVA-C), sweet drinks (NOVA-S), and other drinks (NOVA-D). In our study, moderate alcohol consumption was associated with increased HDL cholesterol [42]. We found a positive association between triglycerides and caffeinated and sweet drinks. Meta-analysis on the effects of coffee consumption on serum lipids previously showed a positive dose-response relationship between coffee intake, cholesterol and triglycerides [43]. Overall, our extension of the NOVA classification to beverages seems to align with already known metabolic links, and the significant associations found between the NOVA processed data and clinical measurements demonstrates the potential of using smartphone food logs for predicting cardiometabolic health.

In a randomised trial, we compared the effects of TRE vs. SDA on metabolic health. We first confirmed compliance with the allocated interventions, thanks to the timestamp and the text annotation of each ingestion event, respectively. We could also confirm limited cross-contamination between both arms, which is a known risk of unblinded trials [44]. Compared to baseline, TRE significantly reduced body weight (−1.6%, SD 2.9), but this reduction in body weight was not significant when compared to SDA. One possible reason for the modest reduction in body weight is that the eating window needs to be shorter to benefit from TRE. Previous studies of TRE reported weight loss with a reduction of eating duration to 10 hours [10,11,15] or even shorter duration (6 hours) [18]. In our study, the eating window was 12-hour long for feasibility reasons: The intervention lasted for 6 months and was trialled in a pragmatic RCT to be applicable to a broader population. Interestingly, rodent studies have shown a dose-response effect in the prevention of obesity with 8-hour TRE leading to maximum benefits, while 12-hour TRE showed fewer benefits [9]. In human studies, a potential dose-response effect is less clear. This may be explained by the fact that rodent studies were done during their entire lifespan, while clinical studies are relatively short compared to the human lifespan. Other explanations of the limited weight loss in our study might be that the clock time of the eating window might play a role in metabolic benefits of TRE [18], or that most other studies were conducted in the US where half of the population have an eating duration > 15h [10], while the population in our study had a mean eating duration below 14h probably for cultural reasons and a lower prevalence of obesity [1]. Future studies should evaluate more stringent forms of TRE (eating duration such as 8 or 10 hours) with longer follow-ups (one year or more) in diverse populations, while thoroughly checking compliance to the intervention.

Finally, using all weight measurements across 5 different visits for each individual showed that some participants seemed to respond well to 12-hour TRE, and the estimated weight loss was significant for 9 of the 25 participants. We found no strong predictors of weight loss and the heterogeneous response to TRE could arise from personal sensitivity to food timing or unknown factors. Further research is needed to reveal the underlying causes.

Our study has several limitations. First, the self-reported ingestion events via a smartphone application could be subject to missing ingestion events and missed photographs. However, the free-text annotation could be added after the meal, if it was socially unacceptable or impossible to take a picture at the time. The timestamp of pictures could not be edited by participants and all smartphones automatically synchronise their date and time online. The chance of mistimed ingestion events was therefore minimal. Second, the unblinded nature of the study and the compliance to the intervention are limitations shared with other trials of dietary interventions [44]. As advised by the CONSORT non-pharmacological trial guidelines, we assessed compliance with the NOVA classification and eating duration, but it’s technically conceivable that one individual willingly limited their pictures of food and drinks only within their assigned TRE window, and not outside of it. Because the motivation of participants was high, we find this unlikely.

Our study shows the potential of mobile technologies to record eating behaviour as part of daily routine and establish relationships with metabolic health. The NOVA classification proved to be a good compromise for summarising nutrition quality. Specifically, annotating ingestion events with NOVA categories remained feasible since a fully quantitative evaluation of eating behaviour is not realistic in large studies. The fact that the NOVA categories were selected in the regression model justifies the approach, and we showed that the inclusion of the smartphone and questionnaire data led to an increase in predictive performance for metabolic health. The randomised trial of TRE vs. SDA confirmed that a TRE intervention is feasible over 6 months in a broad population, but probably requires a shorter eating window for increased benefits on weight loss and other MS components.

## Supporting information

CONSORT NPT checklist

## Data Availability

The data presented in this study are available upon reasonable request to the corresponding author. The data are not publicly available due to confidentiality reasons.

## Supplementary Materials

**Table S1.** Limited overlap of different food and drink categories.

| Categories | NOVA1 (%) | NOVA2 (%) | NOVA3 (%) | NOVA4 (%) | NOVA-A (%) | NOVA-C (%) | NOVA-S (%) | NOVA-D (%) |
| --- | --- | --- | --- | --- | --- | --- | --- | --- |
| NOVA1 | 100 | 0.1 | 0.4 | 0.2 | 0 | 28 | 0 | 7.6 |
| NOVA2 | 0.3 | 100 | 0.7 | 0.1 | 0 | 7.7 | 6.9 | 0 |
| NOVA3 | 1.0 | 0.5 | 100 | 0.3 | 19 | 0 | 0.3 | 0.4 |
| NOVA4 | 0.4 | 0.1 | 0.2 | 100 | 1.6 | 6.9 | 9.7 | 1.7 |
| NOVA-A | 0 | 0 | 90 | 9.7 | 100 | 0 | 6.8 | 0 |
| NOVA-C | 83 | 7 | 0 | 11 | 0 | 100 | 9 | 0 |
| NOVA-S | 0.2 | 28 | 1.9 | 70 | 8.1 | 40 | 100 | 0 |
| NOVA-D | 87 | 0.1 | 2.1 | 10 | 0 | 0 | 0 | 100 |

*Footnotes: For each row, all ingestion events are filtered according to the category in the row name. Each column value represents the percentage of other categories that are present after filtering on the row name. Categories follow and extend the NOVA classification [22–24]: NOVA1: “Unprocessed or minimally processed foods,” NOVA2: “Processed culinary ingredients,” NOVA3: “Processed foods,” NOVA4: “Ultra-processed foods,” NOVA-A: “Alcohol-containing drinks,” NOVA-C: “Caffeinated drinks,” NOVA-S: “Sweet drinks,” NOVA-D: “Other drinks.”*

**Table S2.** Effects of TRE and SDA on eating behaviour.

|  | Time-restricted eating (TRE) |  |  | Standard dietary advice (SDA) |  |  | Between-group difference (95% confidence interval)<sup>c</sup> | P for comparison of pre-post changes between TRE and SDA<sup>b</sup> |
| --- | --- | --- | --- | --- | --- | --- | --- | --- |
| Mean (SD)<sup>a</sup> | Pre-intervention | Post-intervention | P for comparison (pre-post)<sup>b</sup> | Pre-intervention | Post-intervention | P for comparison (pre-post)<sup>b</sup> |  |  |
| NOVA1, % | 43.5 (13.0) | 41.9 (13.9) | 0.35 | 37.8 (12.5) | 44.8 (13.2) | < 0.01 | -8.6 (-14.1; -3.2) | < 0.01 |
| NOVA2, % | 14.7 (6.0) | 15.8 (6.4) | 0.37 | 12.4 (5.8) | 13.9 (5.3) | 0.09 | -0.5 (-3.3; 2.4) | 0.76 |
| NOVA3, % | 17.5 (7.9) | 19.3 (8.7) | 0.10 | 18.0 (7.1) | 17.1 (5.4) | 0.40 | 2.7 (-0.2; 5.6) | 0.08 |
| NOVA4, % | 24.2 (11.1) | 23.0 (9.0) | 0.48 | 31.9 (11.4) | 24.2 (11.8) | < 0.01 | 6.4 (0.4; 12.4) | 0.04 |
| NOVA-A, % | 4.4 (5.3) | 3.9 (4.8) | 0.17 | 4.4 (4.5) | 3.2 (3.4) | < 0.01 | 0.6 (-0.5; 1.7) | 0.27 |
| NOVA-C, % | 19.6 (11.4) | 17.9 (11.7) | 0.28 | 17.4 (6.8) | 13.9 (8.7) | 0.07 | 1.8 (-2.8; 6.5) | 0.44 |
| NOVA-S, % | 4.5 (7.3) | 3.0 (4.6) | 0.08 | 4.4 (4.7) | 4.6 (6.8) | 0.74 | -1.8 (-3.9; 0.3) | 0.12 |
| NOVA-D, % | 3.9 (4.3) | 2.2 (2.5) | 0.02 | 3.9 (3.2) | 3.8 (3.5) | 0.85 | -1.6 (-3.2; -0.0) | 0.06 |
| Eating duration, hours<sup>d</sup> | 15.48 (1.14) | 12.50 (1.29) | < 0.01 | 15.17 (0.96) | 14.89 (1.21) | 0.16 | -2.70 (-3.47; -1.93) | < 0.01 |
| Eating start, hours<sup>d</sup> | 6.83 (0.67) | 8.36 (1.00) | < 0.01 | 6.96 (0.99) | 6.86 (1.08) | 0.40 | 1.63 (1.12; 2.13) | < 0.01 |
| Eating midpoint, hours<sup>d</sup> | 13.84 (1.44) | 13.64 (1.31) | 0.59 | 13.88 (1.18) | 13.05 (0.88) | < 0.01 | 0.62 (-0.24; 1.47) | 0.18 |
| Eating end, hours<sup>d</sup> | 22.31 (1.17) | 20.86 (1.03) | < 0.01 | 22.13 (0.95) | 21.76 (1.10) | 0.01 | -1.07 (-1.62, -0.53) | < 0.01 |

Footnotes: <sup>a</sup> Categories follow and extend the NOVA classification [22–24]: NOVA1: “Unprocessed or minimally processed foods,” NOVA2: “Processed culinary ingredients,” NOVA3: “Processed foods,” NOVA4: “Ultra-processed foods,” NOVA-A: “Alcohol-containing drinks,” NOVA-C: “Caffeinated drinks,” NOVA-S: “Sweet drinks,” NOVA-D: “Other drinks.” <sup>b</sup> All *p* for comparisons were calculated with the Student *t*-test. <sup>c</sup> Mean difference comparing pre-post change between the TRE and SDA groups. <sup>d</sup> Eating duration, the start of the eating interval, the median of the eating interval, and the end of the eating interval, presented as fractional hours, i.e. 1.33h = 1 hour and 20 minutes.

**Table S3.** Baseline characteristics by randomisation group.

|  | **SDA**<br>**(n = 26)** | **TRE**<br>**(n = 28)** | **Both arms**<br>**(n = 54)** |
| --- | --- | --- | --- |
| **Demographics** |  |  |  |
| Age, mean (SD) | 42.5 (14.0) | 44.3 (12.8) | 43.4 (13.3) |
| Marital status<sup>a</sup>, n (%) |  |  |  |
| Married, partnership | 17 (68%) | 23 (92%) | 40 (80%) |
| Single, separated, widow-er | 8 (32%) | 2 (8%) | 10 (20%) |
| Education, n (%) |  |  |  |
| Primary, secondary | 4 (15%) | 2 (7%) | 6 (11%) |
| Apprenticeship, vocational | 5 (19%) | 7 (25%) | 12 (22%) |
| Professional school | 2 (8%) | 5 (18%) | 7 (13%) |
| University, or higher degree | 15 (58%) | 14 (50%) | 29 (54%) |
| Born in Switzerland, n (%) | 21 (81%) | 19 (68%) | 40 (74%) |
| **Clinical characteristics** |  |  |  |
| Weight, kg, median (IQR) | 75.2 (67.9–83.6) | 79.3 (68.5–85.2) | 77.5 (68.0–84.7) |
| BMI, kg/m<sup>2</sup>, median (IQR) | 27.7 (23.8–30.3) | 28.3 (25.2–31.6) | 28.3 (24.6–30.5) |
| WC, cm, median (IQR) | 87.5 (81.0–100.0) | 92.0 (86.0–99.0) | 90.3 (84.0–100.0) |
| Current smokers, n (%) | 2 (8%) | 3 (11%) | 5 (9%) |
| Cardiovascular disease<sup>b</sup>, n (%) | 1 (4%) | 2 (7%) | 3 (6%) |
| Endocrine disorder<sup>c</sup>, n (%) | 1 (4%) | 2 (7%) | 3 (6%) |
| Respiratory disorder, n (%) | 4 (15%) | 5 (18%) | 9 (17%) |
| **Metabolic syndrome (MS)<sup>d</sup>** |  |  |  |
| MS components, n (%) |  |  |  |
| 1. Central obesity | 20 (77%) | 23 (82%) | 43 (80%) |
| 2. Hypertension | 12 (46%) | 11 (39%) | 23 (43%) |
| 3. Impaired fasting glucose | 6 (23%) | 6 (21%) | 12 (22%) |
| 4. High triglycerides | 6 (23%) | 6 (21%) | 12 (22%) |
| 5. Low HDL-cholesterol | 7 (27%) | 8 (29%) | 15 (28%) |
| MS by the IDF definition, n (%) | 6 (23%) | 7 (25%) | 13 (24%) |
| **Lifestyle** |  |  |  |
| Sleep duration<sup>e</sup>, hours, mean (SD) | 7.15 (1.01) | 7.23 (0.93) | 7.19 (0.96) |
| Sleep quality<sup>f</sup>, mean (SD) | 5.1 (2.4) | 5.4 (2.8) | 5.2 (2.6) |
| Physical activity<sup>g</sup>, median (IQR) | 1053 (678–2226) | 1346 (570–1907) | 1191 (594–1920) |

**Figure S1.**
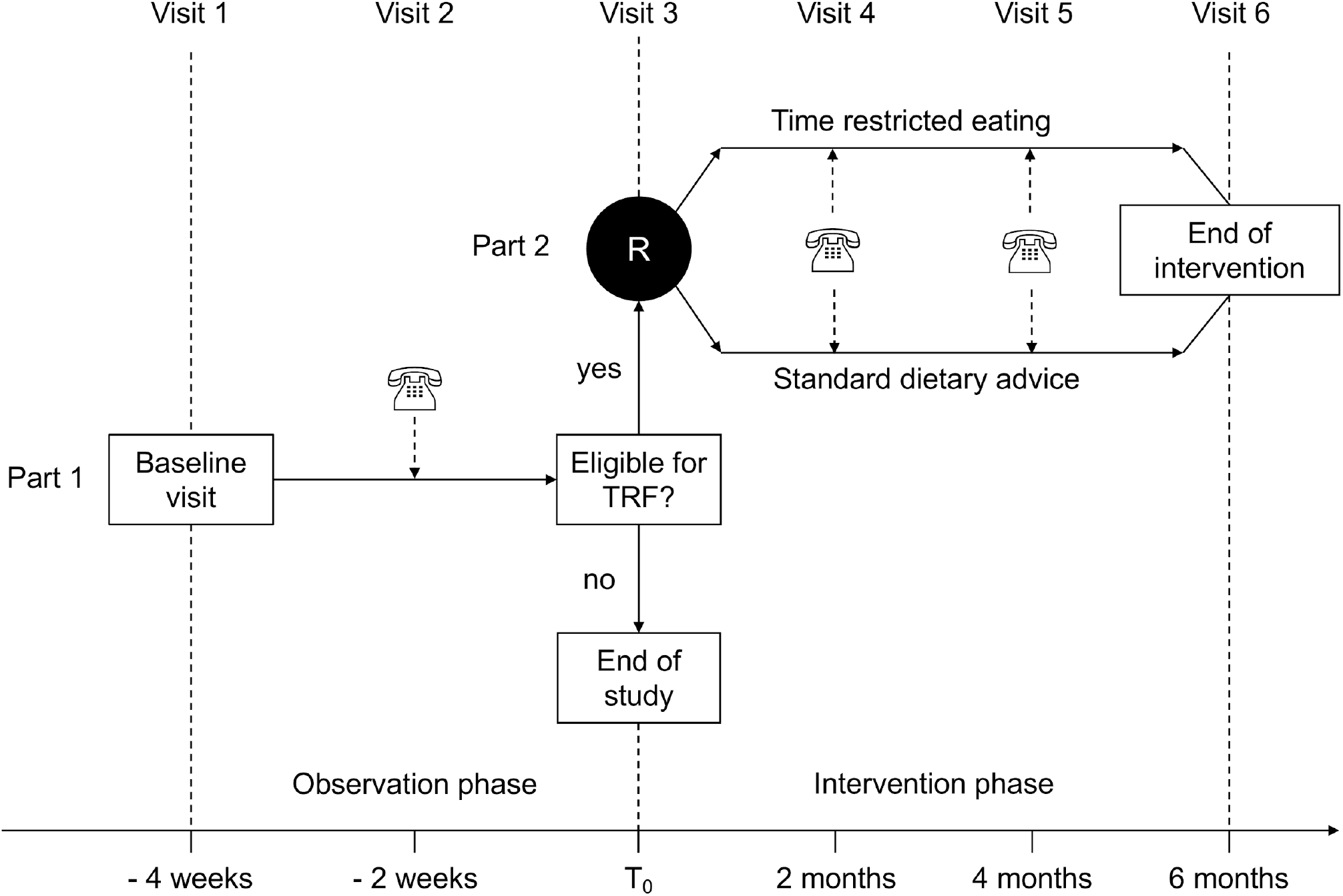
Study design. <u>Legend:</u> Related to the Methods, section 2.1. Study design and flow of participants through the 4-week observation phase, followed by a 6-month intervention phase if they were eligible (see text). In addition to the 3 encounters in person (visits 1, 3, and 6), interim contacts were made over the phone or via email to ensure compliance to the study protocol (visit 2) and record weight measured at home (visits 4 and 5).

**Figure S2.**
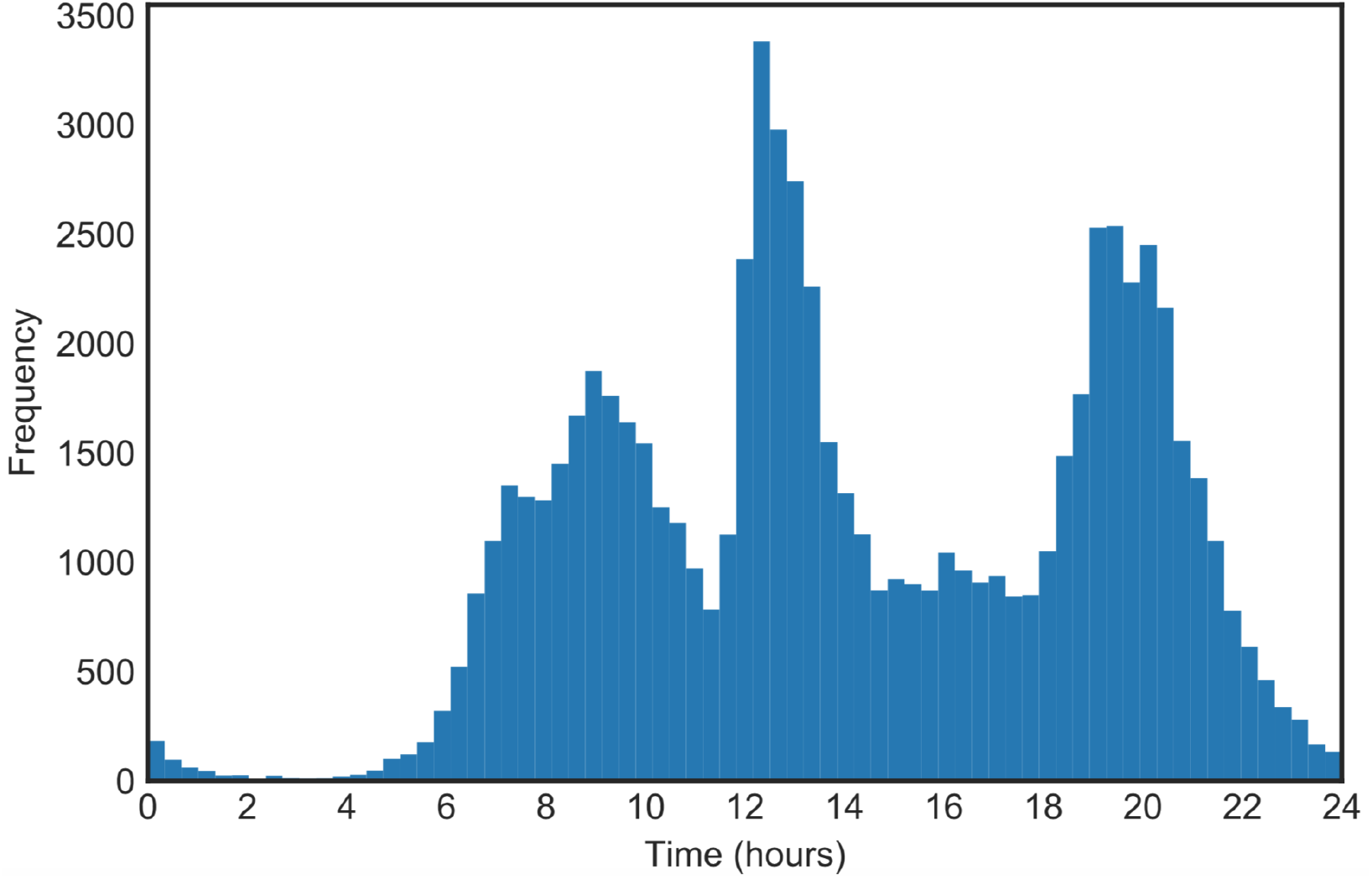
The time profile of ingestion events. <u>Legend:</u> Related to the Methods, section 2.2. The number of recorded ingestion events across all participants as a function of clock time.

**Figure S3.**
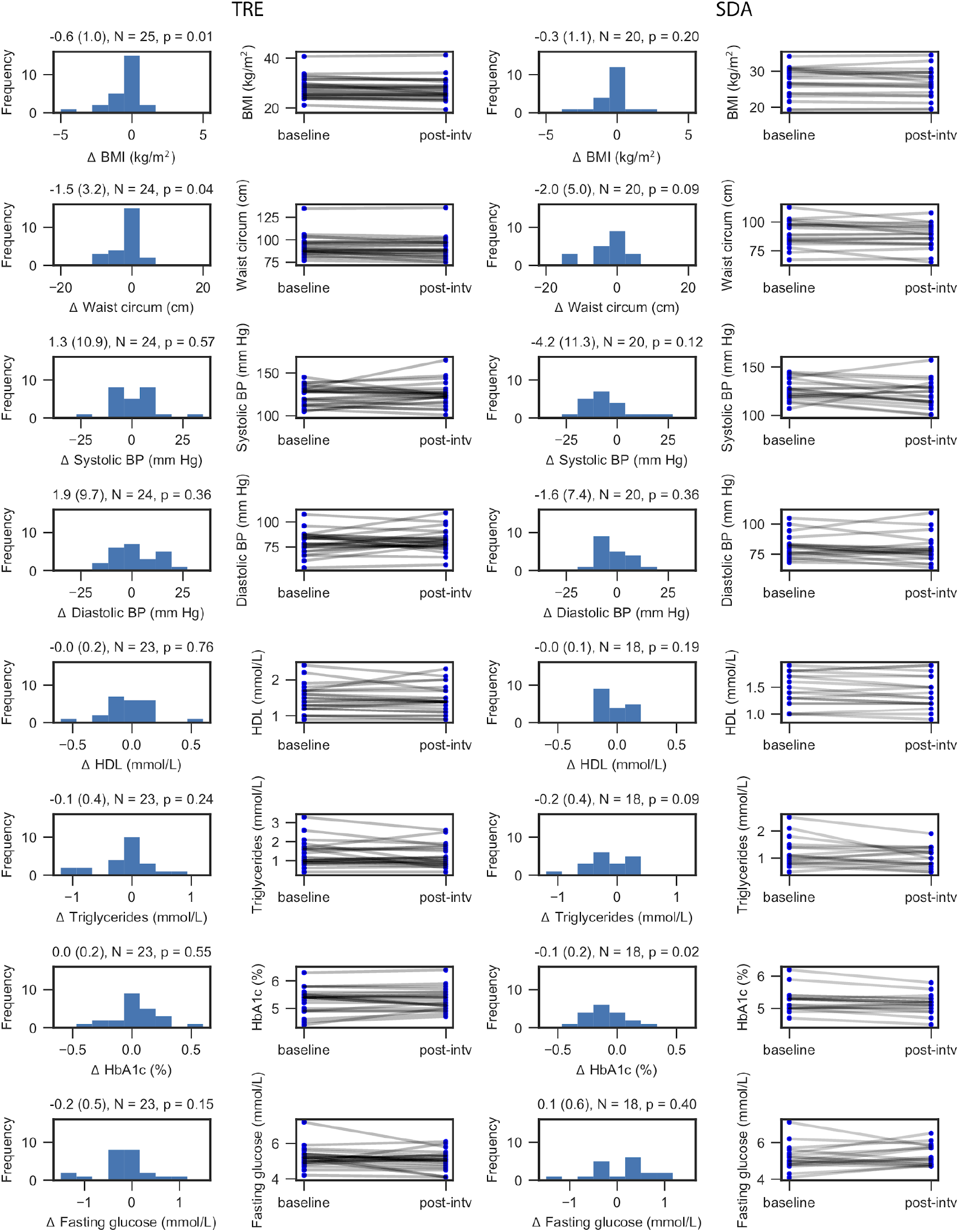
Changes in secondary clinical outcomes after TRE and SDA interventions. <u>Legend:</u> Related to Figure 4. Changes in body mass index (BMI), waist circumference, systolic and diastolic blood pressure (BP), HDL cholesterol, triglycerides, glycated haemoglobin (HbA1c) and fasting plasma glucose after time-restricted eating (TRE, left columns) and standard dietary advice (SDA, right columns). Data are presented as histograms (changes in % from baseline) and individual trajectories pre-/post-intervention. Numbers above each histogram represent the changes (unit on the x-axis) as mean, standard deviation, number of measurements and p-value.

**Figure S4.**
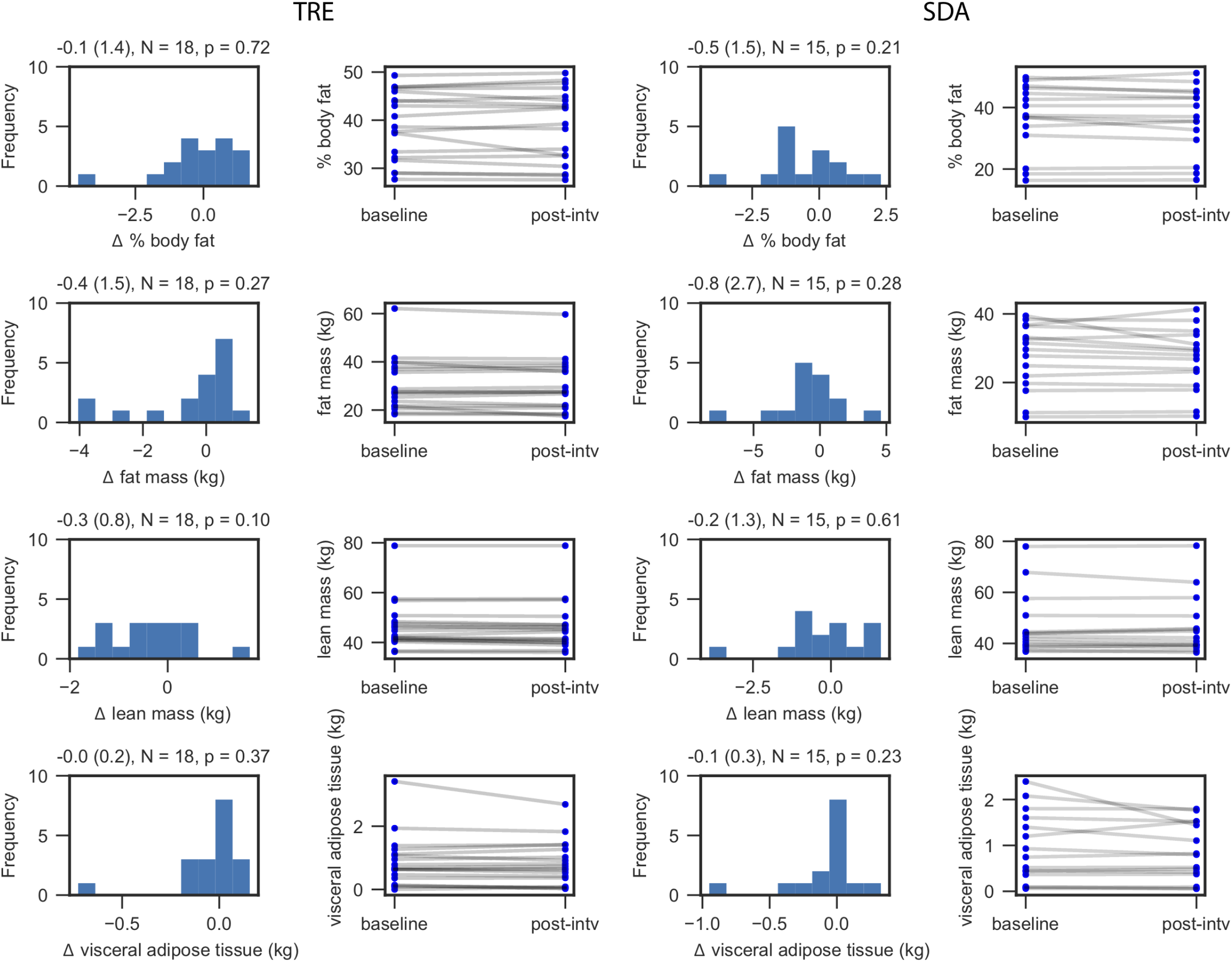
Changes in body composition after TRE and SDA interventions. <u>Legend:</u> Related to Figure 4. Changes in body composition by dual energy X-ray absorptiometry, expressed as the body fat percentage (in % points), fat mass (in kg), lean mass (in kg), and visceral adipose tissue (in kg), after time-restricted eating (TRE, left columns) and standard dietary advice (SDA, right columns). Data are presented as histograms (changes in % from baseline) and individual trajectories pre-/post-intervention. Numbers above each histogram represent the changes (unit on the x-axis) as mean, standard deviation, number of measurements and p-value.

## Author Contributions (CRediT taxonomy)

Conceptualisation, S.P., T.H.C.; methodology, N.E.P., J.M., N.S., F.N., T.H.C.; software, N.E.P., E.N.C.M., S.P., T.H.C.; data collection, J.M., N.S., S.B., G.O., A.G.J., S.U., D.A., D.H., T.H.C.; writing—original draft preparation, N.E.P., T.H.C.; writing—review and editing, all; visualisation, N.E.P., E.N.C.M., S.P., F.N., T.H.C.; supervision, N.S., F.N., T.H.C.; project administration, N.S., T.H.C.; funding acquisition, N.E.P., F.N., T.H.C. All authors have read and agreed to the published version of the manuscript.

## Institutional Review Board Statement

The study was conducted according to the guidelines of the Declaration of Helsinki and the ICH E6 Good Clinical Practice. It was approved by the Ethics Committees of Vaud canton and Bern canton, Switzerland (protocol no 2017-00487).

## Informed Consent Statement

Informed consent was obtained from all participants involved in the study.

## Acknowledgments

The authors wish to thank all the participants to the study and the clinical teams who referred them to the study.

