## Supplementary material for "The effects of time-restricted eating vs. standard dietary advice on weight, metabolic health and the consumption of processed food: A pragmatic randomised controlled trial in community-based adults": CONSORT NPT checklist

2017 CONSORT checklist of information to include when reporting a randomized trial assessing nonpharmacologic treatments (NPTs)*. Modifications of the extension appear in italics and blue.

| Section/Topic Item | Checklist item no. | CONSORT item | Extension for NPT trials | Reported on section / page no. |
| --- | --- | --- | --- | --- |
| Title and abstract |  |  |  |  |
|  | 1a | Identification as a randomized trial in the title |  | page 1 |
|  | 1b | Structured summary of trial design, methods, results, and conclusions (for specific guidance see CONSORT for abstracts) | *Refer to CONSORT extension for abstracts for NPT trials* | page 2 |
| Introduction |  |  |  |  |
| Background and objectives | 2a | Scientific background and explanation of rationale |  | section 1, page 3 |
|  | 2b | Specific objectives or hypotheses |  | section 1, page 4 |
| Methods |  |  |  |  |
| Trial design | 3a | Description of trial design (such as parallel, factorial) including allocation ratio | When applicable, how care providers were allocated to each trial group | section 2-2.1, page 4 |
|  | 3b | Important changes to methods after trial commencement (such as eligibility criteria), with reasons |  | section 2.1, page 5 |
| Participants | 4a | Eligibility criteria for participants | When applicable, eligibility criteria for centers and for *care providers* | section 2.1, page 4-5 |
|  | 4b | Settings and locations where the data were collected |  | section 2.1, page 4 |
| Interventions*†* | 5 | The interventions for each group with sufficient details to allow replication, including how and when they were actually administered | Precise details of both the experimental treatment and comparator | section 2.1, page 4 |
|  | 5a |  | Description of the different components of the interventions and, when applicable, description of the procedure for tailoring the interventions to individual participants. | section 2.1, page 4 |
|  | 5b |  | Details *of whether and* how the interventions were standardized. | yes, section 2.1, page 4 |
|  | 5c. |  | Details *of whether and* how adherence of care providers to the protocol was assessed or enhanced | careful training at start, no assessment during the study |
|  | 5d |  | *Details of whether and how adherence of participants to interventions was assessed or enhanced* | yes, section 2.4, page 6-7 + section 3.4, page 10-11 + Figure 3 |
| Outcomes | 6a | Completely defined pre-specified primary and secondary outcome measures, including how and when they were assessed |  | section 2.5, page 7 |
|  | 6b | Any changes to trial outcomes after the trial commenced, with reasons |  | section 2.1, page 5 |
| Sample size | 7a | How sample size was determined | When applicable, details of whether and how the clustering by care providers or centers was addressed | section 2.6, page 8 |
|  | 7b | When applicable, explanation of any interim analyses and stopping guidelines |  | no interim analyses |
| Randomization: |  |  |  |  |
| - Sequence generation | 8a | Method used to generate the random allocation sequence |  | section 2.1, page 4 |
|  | 8b | Type of randomization; details of any restriction (such as blocking and block size) |  | blocks of 8 by sex, section 2.1, page 4 |
| - Allocation concealment mechanism | 9 | Mechanism used to implement the random allocation sequence (such as sequentially numbered containers), describing any steps taken to conceal the sequence until interventions were assigned |  | computer-generated and concealed until the allocation, section 2.1, page 4 |
| - Implementation | 10 | Who generated the random allocation sequence, who enrolled participants, and who assigned participants to interventions |  | computer-generated, section 2.1, page 4 |
| Blinding | 11a | If done, who was blinded after assignment to interventions (for example, participants, care providers, those assessing outcomes) and how | ~~Whether or not those administering co-interventions were blinded to group assignment~~  If done, who was blinded after assignment to interventions (e.g., participants, care providers, *those administering co-interventions,* those assessing outcomes) and how | no blinding due to the nature of the tested interventions |
|  | 11b | If relevant, description of the similarity of interventions | ~~If blinded, method of blinding and description of the similarity of interventions~~ | same number and duration of visits in the 2 arms |
|  | 11c |  | *If blinding was not possible, description of any attempts to limit bias* | random allocation, section 2.1, page 4 |
| Statistical methods | 12a | Statistical methods used to compare groups for primary and secondary outcomes | When applicable, details of whether and how the clustering by care providers or centers was addressed | section 2.6, page 7-8 |
|  | 12b | Methods for additional analyses, such as subgroup analyses and adjusted analyses |  | section 2.6, page 8-9 |
| Results |  |  |  |  |
| Participant flow (a diagram is strongly recommended) | 13a | For each group, the numbers of participants who were randomly assigned, received intended treatment, and were analyzed for the primary outcome | The number of care providers or centers performing the intervention in each group and the number of patients treated by each care provider or in each center | section 3.1, page 9  + Figure 1 |
|  | 13b | For each group, losses and exclusions after randomization, together with reasons |  | Figure 1 |
|  | 13c |  | *For each group, the delay between randomization and the initiation of the intervention* | no delay |
|  | new |  | Details of the experimental treatment and comparator as they were implemented | section 2.1, page 4 |
| Recruitment | 14a | Dates defining the periods of recruitment and follow-up |  | start in Sept. 2017, end of 6-month follow-up in March 2020 |
|  | 14b | Why the trial ended or was stopped |  | planned end of trial |
| Baseline data | 15 | A table showing baseline demographic and clinical characteristics for each group | When applicable, a description of care providers (case volume, qualification, expertise, etc.) and centers (volume) in each group. | section 3.2, page 9  + Table 1 |
| Numbers analyzed | 16 | For each group, number of participants (denominator) included in each analysis and whether the analysis was by original assigned groups |  | all analyses with intention-to-treat approach (section 2.6), numbers specified in table footnotes and text |
| Outcomes and estimation | 17a | For each primary and secondary outcome, results for each group, and the estimated effect size and its precision (such as 95% confidence interval) |  | section 3.5, page 11  + Figure 4 + Table 2 |
|  | 17b | For binary outcomes, presentation of both absolute and relative effect sizes is recommended |  | n/a |
| Ancillary analyses | 18 | Results of any other analyses performed, including subgroup analyses and adjusted analyses, distinguishing pre-specified from exploratory |  | section 3.6, page 11 + Figure 5 are exploratory |
| Harms | 19 | All important harms or unintended effects in each group (for specific guidance see CONSORT for harms) |  | n/a |
| **Discussion** |  |  |  |  |
| Limitations | 20 | Trial limitations, addressing sources of potential bias, imprecision, and, if relevant, multiplicity of analyses | In addition, take into account the choice of the comparator, lack of or partial blinding, and unequal expertise of care providers or centers in each group | section 4, page 13 |
| Generalizability | 21 | Generalizability (external validity, applicability) of the trial findings | Generalizability (external validity) of the trial findings according to the intervention, comparators, patients, and care providers and centers involved in the trial | section 4, page 13-14 |
| Interpretation | 22 | Interpretation consistent with results, balancing benefits and harms, and considering other relevant evidence |  | section 4, page 13 |
| Other information |  |  |  |  |
| Registration | 23 | Registration number and name of trial registry |  | section 2, page 4 |
| Protocol | 24 | Where the full trial protocol can be accessed, if available |  | upon request to the corresponding author |
| Funding | 25 | Sources of funding and other support (such as supply of drugs), role of funders |  | page 33 |

**Additions or modifications to the 2010 CONSORT checklist. CONSORT = Consolidated Standards of Reporting Trials*

*†The items 5, 5a, 5b, 5c, 5d are consistent with the Template for Intervention Description and Replication (TIDieR) checklist*
